# Simple risk scores to predict hospitalization or death in outpatients with COVID-19 including the Omicron variant

**DOI:** 10.1101/2022.01.14.22269295

**Authors:** Mark H. Ebell, Roya Hamadani, Autumn Kieber-Emmons

**Author notes:** **Corresponding author:** Mark H. Ebell MD, MS, 125 Miller Hall, UGA Health Sciences Campus, Athens, GA 30602.

## Abstract

**Importance:** Outpatient physicians need guidance to support their clinical decisions regarding management of patients with COVID-19, specifically whether to hospitalize a patient or if managed as an outpatient, how closely to follow them.

**Objective:** To develop and prospectively validate a clinical prediction rule to predict the likelihood of hospitalization for outpatients with COVID-19 that does not require laboratory testing or imaging, including during the current Omicron wave.

**Design:** Derivation and temporal validation of a clinical prediction rule, and prospective validation of two externally derived clinical prediction rules.

**Setting:** Primary and urgent care clinics in a Pennsylvania health system.

**Participants:** Patients 12 years and older presenting to outpatient clinics who had a positive polymerase chain reaction test for COVID-19.

**Main outcomes and measures:** Classification accuracy (percentage in each risk group hospitalized) and area under the receiver operating characteristic curve (AUC).

**Results:** Overall, 4.0% of outpatients in the early derivation cohort (5843 patients presenting before 3/1/21), 4.2% in the late validation cohort (3806 patients presenting 3/1/21 to 9/30/21), and 1.9% in an Omicron cohort were ultimately hospitalized. We developed and temporally validated four simple risk scores. The base score included age, dyspnea, and the presence of a comorbidity, with the other scores adding fever, respiratory rate and/or oxygen saturation. All had very good overall accuracy (AUC 0.85-0.87) and classified at least half of patients into a low risk with a < 1% likelihood of hospitalization. Hospitalization rates in the Omicron cohort were 0.22%, 1.3% and 8.7% for the base score. Two externally derived risk scores identified more low risk patients, but with a higher overall risk of hospitalization than our novel risk scores.

**Conclusions and relevance:** A simple risk score applicable to outpatient and telehealth settings can classify over half of COVID-19 outpatients into a very low risk group with a 0.22% hospitalization risk in the Omicron cohort. The Lehigh Outpatient COVID Hospitalization (LOCH) risk score is available online as a free app: https://ebell-projects.shinyapps.io/LehighRiskScore/.

**Key points:** *Question:* Is it possible to predict the eventual likelihood of hospitalization for outpatients with COVID-19 using simple non-laboratory based risk scores?

*Findings:* We created and temporally validated in the same population 4 risk scores with 3 to 5 predictors that do not require laboratory testing. Groups with low (0.34% to 0.89%), moderate (4.0% to 6.2%), and high-risk (19.2% to 25.2%) of hospitalization were identified. The risk scores were also accurate in an Omicron dominant cohort with hospitalization rates of 0.22% to 0.43% in the low-risk groups, 1.3% to 1.7% in the moderate risk groups, and 8.7% to 15.3% in the high risk groups.

*Meaning:* Simple risk scores can help support decisions about hospitalization in the outpatient setting.

## Introduction

Most patients with COVID-19 are initially evaluated in the outpatient setting, and a decision must be made whether to manage them as outpatients or to hospitalize them. For those managed initially in the outpatient setting, a decision must also be made whether they require close follow-up or whether they are at a low risk of deterioration and can be told to simply follow-up as needed. A risk score designed for outpatients could help support clinician decision-making around hospitalization and follow-up.

However, while many COVID-19 risk scores have been proposed, almost all have been developed and validated to predict mortality in hospitalized patients.^1^ These include the 4C risk score,^2^ the ABCS risk score,^3^ and the COVID-GRAM risk score.^4^ Almost all of these risk scores require laboratory tests and in some cases imaging, which is often unavailable in the outpatient or telehealth settings. An exception is the COVID-NoLab risk score for inpatient mortality, which requires only oxygen saturation, age, and the respiratory rate.^5^ Additionally, the OutCoV score was derived and internally validated in Switzerland to predict the likelihood of hospitalization among outpatients and does not require laboratory testing.^6^ It consists of 5 easily ascertained variables: age, fever, dyspnea, hypertension, and chronic respiratory disease. However, it has not been externally validated.

We assembled a dataset of consecutive patients who had been diagnosed with COVID-19 in an outpatient primary care or urgent care clinic in a large health system in Pennsylvania. The primary objective was to develop and temporally validate simple risk scores that do not require laboratory tests or imaging to predict the likelihood of hospitalization or death in outpatients with COVID-19. A secondary objective was to prospectively validate two previously developed mortality risk scores that did not require laboratory testing, the OutCoV score developed in outpatients and the COVID-NoLab risk scores developed originally for inpatients.^5,6^

## Methods

### Population studied

Lehigh Valley Hospital, Inc. (LVH) is a not-for-profit academic community hospital, which is a legal entity of Lehigh Valley Health Network (LVHN). LVHN has grown to become the largest health care provider in the Pennsylvania Lehigh Valley region, with nine not-for-profit hospital campuses, nearly 2,000 inpatient beds, more than 275 physician practice locations, 25 health centers and 20 ExpressCARE locations, and includes the region’s only Children’s Hospital. The LVH hub is located in the Lehigh Valley region of eastern Pennsylvania, approximately 90 miles from New York City, 60 miles from Philadelphia, and 20 miles from the New Jersey border, and encompasses Allentown, the third largest city in Pennsylvania. According to the 2020 Census, Allentown is home to 125,845 people. Of this population, 52% identify as Latino, 14.7% as Black/African American, and 2.9% as Asian. ^7^

The electronic health record was used to identify all outpatients diagnosed with COVID-19 by polymerase chain reaction (PCR) test between March 13, 2020 and September 30, 2021. Outpatients with an outpatient PCR diagnosis of COVID-19 from December 20, 2021 to January 7, 2022 were additionally identified to prospectively validate the risk score in patients during the Omicron phase of the pandemic. Data were obtained from query of LVHN’s Epic electronic medical record and included all primary care and ExpressCARE (urgent care) outpatient visits during the time window that included a COVID-19 PCR order from that visit, and a subsequent positive COVID-19 PCR test result. Patients with missing data for age, respiratory rate, or oxygen saturation were excluded from the population used to derive and internally validate the clinical prediction rules, as were patients less than 12 years of age.

### Analytic plan

Univariate analysis was performed using a chi-square test for dichotomous variables and Student’s t-test for normally distributed continuous variables to identify individual patient characteristics associated with hospitalization or death. Potential cutpoints for continuous variables were selected by inspection of histograms and contingency tables. All comorbidities associated with hospitalization were combined into a single “any comorbidity” variable. Patients with hospitalization within the study period but prior to the date of the outpatient visit were excluded.

Patients presenting prior to March 1, 2021 were used to derive the risk score (early cohort), and patients presenting from March 1, 2021 onwards were used to validate it (late cohort). The risk score was also validated in a cohort presenting when Omicron was the predominant variant, with patients presenting between December 20, 2021 to January 7, 2022. That end date was chosen to assure at least 2 weeks of follow-up for all patients. Logistic regression was performed with hospitalization or death as the dependent variable, and all patient characteristics significantly associated with hospitalization or death in univariate analysis as the independent variables. We evaluated multiple models using different cutoffs for oxygen saturation, age and respiratory rate in the derivation group to identify the models with the highest area under the receiver operating characteristic curve.

The final models were converted to simple point scores by dividing each beta-coefficient by the smallest beta-coefficient and rounding the resulting number. Low, moderate and high-risk groups were identified by inspection of a table showing the likelihood of hospitalization associated with each risk score. The primary goal was to identify as large as possible a low-risk group with a less than 1% likelihood of hospitalization. The resulting risk scores were then validated in the late cohort of outpatients and also in the Omicron cohort. Analysis was performed using Stata v. 17 (StataCorp, College Station, Texas). The study was approved by the University of Georgia Human Subjects Office (PROJECT00004060) and LVHN staff received approval to use the University of Georgia determination.

## Results

Data were available for 13,418 outpatients diagnosed with COVID-19 between March 20, 2020 and September 30, 2021. After excluding patients under age 12 and patients with missing data, the final dataset used to derive and internally validated the risk scores included 9649 outpatients with COVID-19. The early cohort had 5843 patients with a 28 day hospitalization rate of 4.0% while the late cohort had 3806 patients with a 28 day hospitalization rate of 4.2%. A total of 641 patients in the entire cohort were hospitalized and of that number, 55 died following their outpatient COVID-19 diagnosis, all of whom were hospitalized. Of the hospitalized patients, 89 of 641 (13.9%) were hospitalized on the same day as their outpatient visit. The characteristics of patients who were hospitalized or died and of those who were not in the derivation and internal validation cohorts are summarized in Table 1. Increasing age, increasing respiratory rate, lower oxygen saturation, a complaint of dyspnea, and all comorbidities were associated with an increased likelihood of hospitalization.

**Table 1.** Characteristics of included patients in the derivation and internal validation cohort.

| Clinical parameter | Non-Hospitalized | Hospitalized | P value |
| --- | --- | --- | --- |
| **Vital signs** |  |  |  |
| Mean age | 41.9 | 60.2 | < 0.001 |
| Age category |  |  |  |
| < 50 years | 5960 | 91 (1.5%) | < 0.001 |
| 50-59 years | 1631 | 84 (4.9%) |  |
| 60-69 years | 1032 | 100 (8.8%) |  |
| 70+ years | 623 | 128 (17.0%) |  |
| Respiratory rate > 30/minute | 3/9246 (0.03%) | 3/403 (0.74%) | < 0.001 |
| Respiratory rate > 20/minute | 261/9246 (2.8%) | 43/403 (10.7%) | < 0.001 |
| Oxygen saturation ≤ 95% | 330/9246 (3.6%) | 111/403 (27.5%) | < 0.001 |
| Temperature ≥ 100.4 F | 433/9246 (4.7%) | 36/403 (8.9%) | 0.008 |
| **Comorbidities** |  |  |  |
| Type 1 or 2 diabetes mellitus | 806/9246 (8.7%) | 142/403 (35.2%) | < 0.001 |
| Asthma | 918/9246 (9.9%) | 51/403 (12.7%) | 0.07 |
| COPD or chronic bronchitis | 192/9246 (2.1%) | 68/403 (16.9%) | < 0.001 |
| Hypertension | 1950/9246 (21.1%) | 257/403 (63.8%) | < 0.001 |
| Cardiovascular disease | 356/9246 (3.9%) | 88/403 (21.8%) | < 0.001 |
| Chronic kidney disease | 151/9246 (1.6%) | 55/403 (13.7%) | < 0.001 |
| Chronic liver disease | 362/9246 (3.9%) | 40/403 (9.9%) | < 0.001 |
| Cancer | 340/9246 (3.7%) | 49/403 (12.2%) | < 0.001 |
| Any of the above comorbidities | 3028/9246 (32.8%) | 308/403 (76.4%) | < 0.001 |
| **Symptom** |  |  |  |
| Dyspnea | 828/9246 (9.0%) | 168/403 (41.7%) | < 0.001 |

Note: For age, percentage shown is percentage within an age category who were hospitalized. For all other parameters it is the percentage of hospitalized and non-hospitalized with that comorbidity or parameter.

The Omicron cohort had 6138 patients with at least 2 weeks of follow-up, and a 1.9% overall rate of hospitalization. As in the earlier cohorts, the median age of unhospitalized patients in the Omicron cohort was 41 years (interquartile range [IQR] 29 to 54 years), while for hospitalized patients it was 63 years (IQR 51 to 73 years).

Four logistic regression models were developed based on the assumption that in telehealth settings temperature, oxygen saturation, respiratory rate, or all three may not be available. Model A used age, dyspnea, and the presence of a comorbidity; Model B added fever; Model C added fever and a respiratory rate > 20/minute; and Model D added fever and oxygen saturation ≤ 95%. A model adding fever, elevated respiratory rate and low oxygen saturation had excessive collinearity for the respiratory rate predictor and is not shown. The models are shown in Table 2, along with corresponding points assigned to each variable based on the beta-coefficient. The four models had very good discrimination based on the area under the receiver operating characteristic curve, with a range from 0.847 to 0.0.872. ROC curves for models A and D are shown in Figure 1.

**Table 2.** Multivariate models, showing assignment of points based on the beta-coefficients

| Variable | β-coefficient | P>z | $\beta$/lowest $\beta$ | Points |
| --- | --- | --- | --- | --- |
| **Model A: Base score, AUROCC = 0.847** |  |  |  |  |
| Dyspnea | 1.3740 | 0.000 | 1.50 | 1.5 |
| Any comorbidity * | 1.1146 | 0.000 | 1.22 | 1 |
| Age 50-59 years | 0.9165 | 0.000 | 1.00 | 1 |
| Age 60-69 years | 1.4885 | 0.000 | 1.62 | 1.5 |
| Age 70+ years | 2.1250 | 0.000 | 2.32 | 2.5 |
| Constant | -5.0277 |  |  |  |
| **Model B: Adds fever, AUROCC = 0.850** |  |  |  |  |
| Dyspnea | 1.3807 | 0.0000 | 1.77 | 2 |
| Any comorbidity * | 1.1059 | 0.0000 | 1.42 | 1.5 |
| Age 50-59 years | 0.9013 | 0.0000 | 1.16 | 1 |
| Age 60-69 years | 1.4902 | 0.0000 | 1.91 | 2 |
| Age 70+ years | 2.1290 | 0.0000 | 2.73 | 3 |
| Temperature $\geq 100.4$ | 0.7798 | 0.0020 | 1.00 | 1 |
| Constant | -5.0753 |  |  |  |
| **Model C: Adds respiratory rate > 20/minute, AUROCC = 0.853** |  |  |  |  |
| Respiratory rate > 20/minute | 0.8615 | 0.000 | 1.11 | 1 |
| Dyspnea | 1.3167 | 0.000 | 1.69 | 1.5 |
| Any comorbidity * | 1.0753 | 0.000 | 1.38 | 1.5 |
| Age 50-59 years | 0.8967 | 0.000 | 1.15 | 1 |
| Age 60-69 years | 1.4709 | 0.000 | 1.89 | 2 |
| Age 70+ years | 2.1074 | 0.000 | 2.71 | 3 |
| Temperature $\geq 100.4$ | 0.7784 | 0.002 | 1.00 | 1 |
| Constant | -5.0856 |  |  |  |
| **Model D: Adds oxygen saturation $\leq 95\%$, AUROCC = 0.872** |  |  |  |  |
| Oxygen saturation $\leq 95\%$ | 1.4937 | 0.000 | 2.37 | 2.5 |
| Dyspnea | 1.2769 | 0.000 | 2.03 | 2 |
| Any comorbidity * | 1.0810 | 0.000 | 1.72 | 1.5 |
| Age 50-59 years | 0.7538 | 0.001 | 1.20 | 1 |
| Age 60-69 years | 1.1124 | 0.000 | 1.77 | 2 |
| Age 70+ years | 1.6574 | 0.000 | 2.63 | 2.5 |
| Temperature $\geq 100.4$ | 0.6295 | 0.018 | 1.00 | 1 |
| Constant | -5.1852 |  |  |  |

\* Type 1 or 2 diabetes mellitus, asthma, COPD or chronic bronchitis, hypertension, cardiovascular disease, chronic kidney disease, chronic liver disease, or cancer

**Figure 1.**
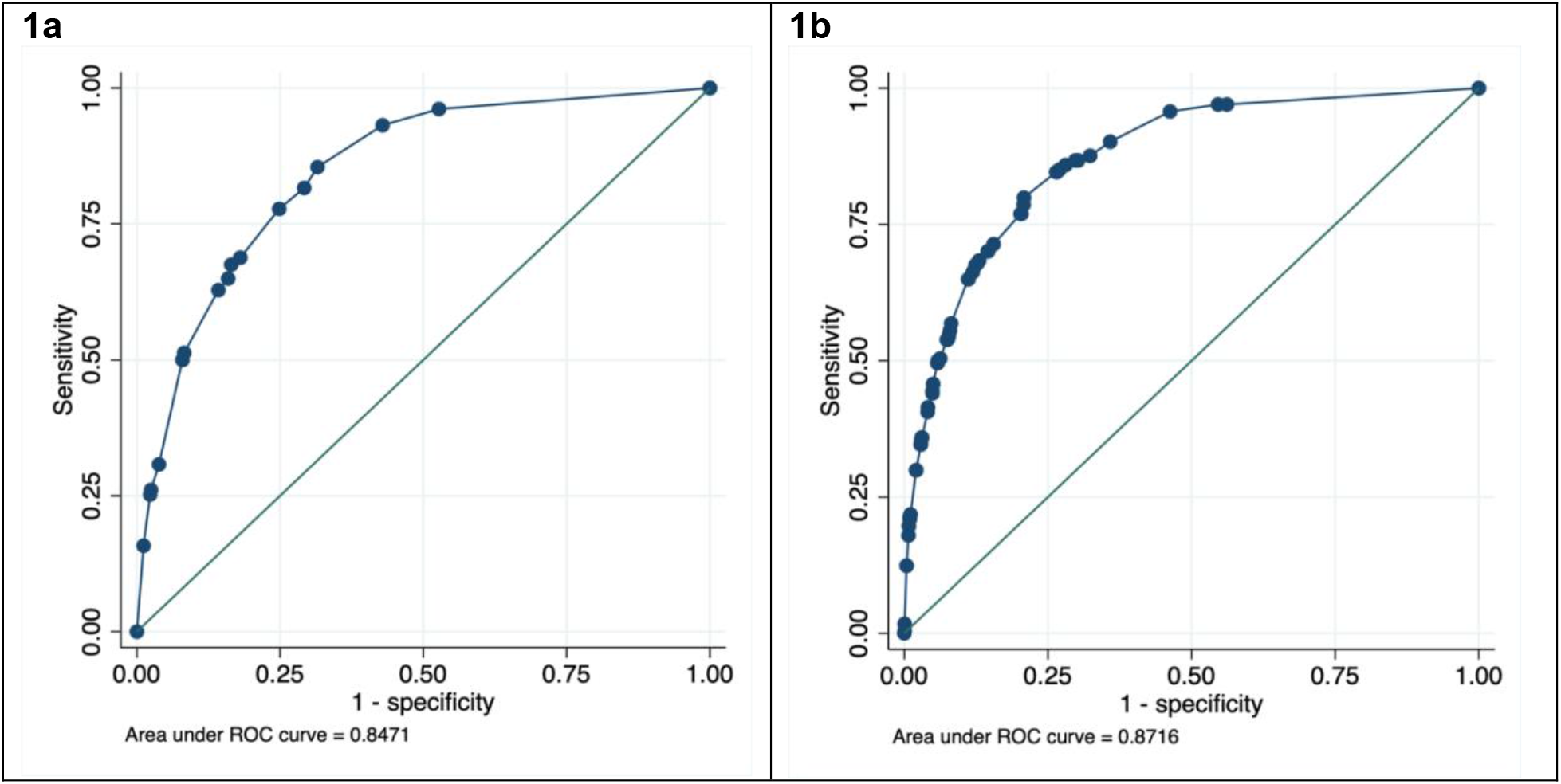
Receiver operating characteristic curves for 1a: Model A (age, comorbidities and dyspnea only) and 1b: Model D (adding fever and oxygen saturation)

The performance of each risk score in the early (derivation), late (validation), and Omicron cohorts is summarized in Table 3. Each model classified at least half of patients in the low-risk group, with a 0.34% to 0.47% risk of hospitalization or death in the early derivation cohort and a 0.79% to 0.89% risk of hospitalization in the later validation cohort.

**Table 3.** Classification accuracy of 3 novel risk scores and 2 externally derived risk scores in the early derivation, late validation, and Omicron cohorts.

|  | Early derivation cohort | Late validation cohort | Omicron cohort |
| --- | --- | --- | --- |
| **Dates of data collection** | **3/20/20-2/28/21** | **3/1/21-9/30/21** | **12/20/21-1/7/22** |
| **Risk group (points)** | **Hospitalized/**<br>**Total (%)** | **Hospitalized/**<br>**Total (%)** | **Hospitalized/**<br>**Total (%)** |
| **Overall** | 233/5839 (4.0%) | 157/3775 (4.2%) | 74/6138 (1.2%)* |
| **Model A risk score** |  |  |  |
| Low (0) | 9/2660 (0.34%) | 15/1849 (0.81%) | 7/3144 (0.22%) |
| Moderate (1.0 – 2.5) | 105/2600 (4.0%) | 84/1623 (5.2%) | 34/2613 (1.3%) |
| High ($\geq 3.0$) | 120/583 (20.6%) | 70/334 (21.0%) | 33/381 (8.7%) |
| **Model B risk score** |  |  |  |
| Low (0 – 1.0) | 15/3188 (0.47%) | 19/2135 (0.89%) | 7/1724 (0.41%) |
| Moderate (1.5 – 3.5) | 96/2035 (4.7%) | 80/1320 (6.1%) | 22/1302 (1.7%) |
| High ($\geq 4.0$) | 123/620 (19.8%) | 70/351 (19.9%) | 35/327 (10.7%) |
| **Model C risk score** |  |  |  |
| Low (0 – 1.0) | 14/3172 (0.44%) | 18/2119 (0.85%) | 7/1621 (0.43%) |
| Moderate (1.5-3.5) | 98/2043 (4.8%) | 82/1328 (6.2%) | 21/1345 (1.6%) |
| High ($\geq 4.0$) | 131/637 (20.6%) | 69/359 (19.2%) | 36/387 (9.3%) |
| **Model D risk score** |  |  |  |
| Low (0 – 1.0) | 10/3205 (0.33%) | 16/2020 (0.79%) | 3/1649 (0.18%) |
| Moderate (1.5- 4.5) | 106/2349 (4.5%) | 79/1481 (5.3%) | 25/1469 (1.7%) |
| High ($\geq 5.0$) | 117/465 (25.2%) | 74/305 (24.3%) | 36/235 (15.3%) |
| **COVID-NoLab risk score** |  |  |  |
| Low (0-1) | 142/3544 (4.0%) | 70/2488 (2.8%) | 12/2081 (0.6%) |
| Moderate (2-5) | 259/2254 (11.5%) | 132/1302 (10.1%) | 45/1253 (3.6%) |
| High ($\geq 6$) | 29/45 (64.4%) | 9/16 (56.3%) | 7/19 (36.8%) |
| **OutCoV risk score** |  |  |  |
| Low ($< 3$) | 163/4340 (3.8%) | 81/2929 (2.8%) | 18/2534 (0.71%) |
| Moderate (3.0 – 5.0) | 220/1392 (15.8%) | 112/818 (13.7%) | 32/769 (4.2%) |
| High ($\geq 5.5$) | 47/111 (42.3%) | 18/59 (30.5%) | 14/50 (28.0%) |

\* For the subset with measured temperature available used for risk scores B-D, there were 64 hospitalizations out of 3353 outpatients (1.9%)

The overall likelihood of hospitalization in the Omicron cohort was 1.2%, lower than in the earlier cohorts. In the Omicron cohort, the hospitalization rates for Risk Score A were 0.22% in the low-risk group, 1.3% in the moderate risk group, and 8.7% in the high-risk group. Again, over 60% of patients were classified in the low-risk group. The other risk scores performed similarly well in the Omicron cohort.

The classification accuracy of the OutCoV and COVID-NoLab risk scores in the late cohort is also shown in Table 3. These risk scores identified more patients in the low-risk groups, but also had higher rates of hospitalization in that group, 3.8% to 4.0% in the early cohort and 2.8% in the late cohort. In the Omicron cohort, both risk scores identified low-risk groups with hospitalization rates of 0.6% to 0.71% but classified only a small number of patients in the high-risk groups.

## Discussion

We developed 4 simple risk scores for hospitalization in outpatients with COVID-19, and prospectively validated them in the same population. The base risk score A included age, dyspnea and the presence of a comorbidity, with risk score B adding fever, risk score C adding fever and the respiratory rate, and risk score D adding fever and the oxygen saturation. All four models all had very good accuracy based on the AUC (0.847 to 0.872).

In the validation group, the risk scores classified at least half of patients being in a low risk group with a likelihood of hospitalization less than 1%. These patients could potentially be managed initially as outpatients with guidance to contact their primary care physician in the event of worsening symptom. The moderate risk groups for the 4 risk scores had a 5.2% to 6.2% likelihood of hospitalization in the late validation cohort, which is similar to that for the population as a whole. These patients could potentially be managed as outpatients initially, but with close follow-up. For example, they could be given an oxygen saturation monitor with daily check-in from a nurse or other clinician to assess their status. Finally, about 9% of patients fell into a high-risk group with a 19.2% to 24.3% likelihood of hospitalization. It may be appropriate to refer these patients to the emergency department for more detailed evaluation including imaging, laboratory testing, and a period of observation.

Perhaps most importantly, we also evaluated the risk scores in a group of patients recruited when the Omicron variant represented over 99% of cases in the United States, from December 20, 2020 to January 21, 2022. The overall hospitalization rate was lower, as has been reported by others. Overall, the risk scores still had good accuracy. The simplest risk score classified approximately half of patients in a very low risk group with only a 0.22% risk of hospitalization.

We also evaluated the accuracy of two previously developed clinical prediction rules. The COVID-NoLab score was originally designed for inpatient risk prediction, but very few patients in our sample had either oxygen saturation < 93% or respiratory rate > 30 breaths/minute. While it identified very few high-risk patients, it classified 62% of outpatients in the Omicron cohort into a low-risk group of whom 0.6% were hospitalized. The OutCoV risk score similarly identified few high-risk patients, but did classify 75% of patients into a low-risk group with a 0.71% risk of hospitalization. Both scores therefore identify more low-risk outpatients than the novel risk scores, but at the cost of a higher risk of hospitalization in those low-risk groups (0.6% to 0.71% versus 0.22%). Depending on the tolerance for risk, this may be an acceptable trade-off.

### Strengths and Limitations

Our study had several notable strengths. We had access to a large, real-world population of outpatients being managed in primary and urgent care settings. The risk scores developed in the earliest 60% of patients validated very well in the later cohort of patients, with similar classification accuracy, and also in a cohort of patients recruited while Omicron was the dominant variant. Most importantly, the risk scores all validated well in a large cohort of patients during a period when Omicron was the dominant variant. The risk scores are simple, and even the simplest had very good overall accuracy and was able to identify clinically useful low, moderate, and high risk groups. While telehealth patients were not included in our sample, all of our clinical variables (especially for Risk Score A) can be easily obtained in that setting.

The study also had limitations. Data were obtained retrospectively from the electronic health record, and we were unable to directly verify diagnoses. Additionally, visits to the LVHN network for COVID-19 concerns were available both through telehealth (video or phone encounters) as well as outpatient and urgent care visits during the initial months of the pandemic and then also during surge windows. Thus, this sample will not include some individuals in the health system who chose to do telehealth visits for their symptoms and COVID-19 testing needs. Selection bias regarding patient characteristics of those individuals who did come to an in-person visit could have affected the risk scores. However, it is more likely that those who sought in-person outpatient or urgent care visits were sicker than the overall universe of COVID-19 patients, and hence the risk score population may be biased towards those with a higher risk of hospitalization. It will also be important to evaluate the risk score in vaccinated versus unvaccinated populations, possibly incorporating lack of vaccination as an additional risk factor. However, we did not have access to full vaccination records, only those administered within the LVHN. Finally, we only had 2 weeks of follow-up for Omicron patients but are continuing to gather data and will update the online risk calculator as it emerges.

## Conclusion

We have developed and temporally validated simple risk scores for outpatients with COVID-19 that identify a large proportion of outpatients who have very low risk of hospitalization (0.22% in the low-risk group for the Omicron cohort). They also classify about one in nine Omicron patients into a high-risk group who have a substantial likelihood of eventual hospitalization and warrant very close follow-up or referral to the emergency department for further evaluation. Risk score A, which we call the Lehigh Outpatient COVID Hospitalization (LOCH) risk score is available online as a simple interactive app: https://ebell-projects.shinyapps.io/LehighRiskScore/.

## Data Availability

All data produced in the present study are available upon reasonable request to the authors.

